# Mortality rates from COVID-19 in Spain and Italy, Lessons for the UK!

**DOI:** 10.1101/2020.05.13.20100453

**Authors:** Adrian Davis, Irene Stratton, Richard Quinton, Gerard O’Donoghue, Graham Roberts

**Affiliations:** University College London, Ear Institute, London, UK And AD CAVE SOLUTIONS Limited, 67 Rossendale Way, London, NW1 0XA, UK; Cheltenham General Hospital, Gloucestershire Retinal Research Group, Cheltenham, Gloucestershire, UK, 07979367684; Newcastle Upon Tyne Hospitals NHS Trust, Newcastle Upon Tyne, Newcastle upon Tyne, UK; NIHR Biomedical Research Centre at Nottingham, Otorhinolaryngology, Nottingham University Hospitals NHS Trust, UK, 07970091724; University College Cork, HRC-Clinical Research Facility, University Hospital Waterford, Dunmore Road, Waterford, Cork, IE

**Keywords:** EPIDEMIOLOGY, PUBLIC HEALTH, INFECTION

## Abstract

**Background:** The Covid-19 pandemic brings major new challenges to health services resulting from the lack of a vaccine and from the enormous resources it can consume over a prolonged period. The available control measures are currently limited to quarantining, contact ‘tracking and tracing’ and social distancing. Disease transmission to health care workers is common and deaths among clinical and nursing staff have been reported in the UK (where serious concerns about the availability of personal protective equipment – PPE – have been raised) and elsewhere; particularly in Lombardy, where General Practitioners (*Medici di Base*) have died in disproportionate numbers.

**Methods:** Data from Italy and Spain was obtained from publicly available sources which are more comprehensive than that available to date (April 2020) in the UK and the advanced timing of the crisis in these locations allows their sources to propose a strategy of allocating staff roles with due respect to the age and sex of staff in order to reduce the pressure on the limited resource of critical care beds and diminish the quantity of hospital acquired infections encountered in treating the known large proportions of patients who were infected as healthcare workers. Such strategy would dramatically reduce unnecessary death and illness in the caring professions and assist employers in reasonable health and safety compliance for their workforce.

In addition to those staff employed in patient care roles prior to the pandemic, doctors and nurses who have recently retired from the NHS have been invited via their respective regulatory bodies such as the United Kingdom General Medical Council (GMC) and Nursing & Midwifery Council (NMC) to return to the front-line. Those who are qualified for clinical and nursing roles, but who are now working in research, education and management roles have also been welcomed by the NHS and placed in clinical roles undertaking face-to face delivery of care. The first COVID-19-related deaths reported in NHS doctors were all male clinicians from BAME background over 50 years of age.

**Results:** Herein, we construct a measure of fatality as a function of population Covid-19 status to examine the data from Spain and Italy, where the pandemic struck much sooner. This shows a doubling in relative risk for men compared to women and a 20-fold increase for those 60-19 compared to women 40-49.

**Conclusion:** The analyses suggest a stratified approach to allocating staff, through making these results available to those who volunteer for front line roles. This should result in preserving as many of the valuable older doctors and nurses as possible with their expertise, experience, managerial skills and to maintain the coherence and leadership of their teams and research groups.

## Introduction

The Covid-19 pandemic brings major new challenges to health services resulting from the lack of a vaccine and from the enormous resources it can consume over a prolonged period. The available control measures are currently limited to quarantining, contact ‘tracking and tracing’ and social distancing. Disease transmission to health care workers is common and deaths among clinical and nursing staff have been reported in the UK (where serious concerns about the availability of personal protective equipment – PPE – have been raised) and elsewhere; particularly in Lombardy, where General Practitioners (*Medici di Base*) have died in disproportionate numbers.

Data from Italy and Spain is more comprehensive than that available to date in the UK and the advanced timing of the crisis in these locations allows their sources to propose a strategy of allocating staff roles with due respect to the age and sex of staff in order to reduce the pressure on the limited resource of critical care beds and diminish the quantity of hospital acquired infections encountered in treating the known large proportions of patients who were infected as healthcare workers. Such stratergy would dramatically reduce unnecessary death and illness in the caring professions and assist employers in reasonable health and safty compliance for their workforce.

In addition to those staff employed in patient care roles prior to the pandemic, doctors and nurses who have recently retired from the NHS have been invited via their respective regulatory bodies such as the United Kingdom General Medical Council (GMC) and Nursing & Midwifery Council (NMC) to return to the front-line. Those who are qualified for clinical and nursing roles, but who are now working in research, education and management roles have also been welcomed by the NHS and placed in clinical roles undertaking face-to face delivery of care. The first COVID-19-related deaths reported in NHS doctors were all male clinicians over 50 years of age.

Herein, we examine the data from Spain and Italy, where the pandemic struck much sooner, and suggest a stratified approach to allocating staff to front line roles, in order to preserve as many of the valuable older doctors and nurses, their expertise, experience, managerial skills and to maintain the coherence and leadership of their teams and research groups.

## Methods

We extracted data on numbers, demographic characteristics and fatality of people who tested positive for COVID-19 from Italy and Spain from their respective Health Ministry websites, and determined fatality outcomes stratified by age and sex.

Data for age, specialty and sex of Italian doctors who have died in the COVID-19 pandemic were extracted from obituary notices (1)

Data on clinical and nursing staff who have died were extracted from the Daily Telegraph All analyses were carried out with SAS v 9.4

## Results

The basic data we extracted are shown in Table 1 and we performed logistic regression using country, sex and age-group as the major independent variables. The analyses show that country, sex and age-group were all highly significant (p<0.001 for each). Table 2 shows the relative risk ratios for the sex and age-group contrasts using women 40-49 as the comparison group (due to the numbers available at that sex and age category to make stable comparisons).

**Table 1.**
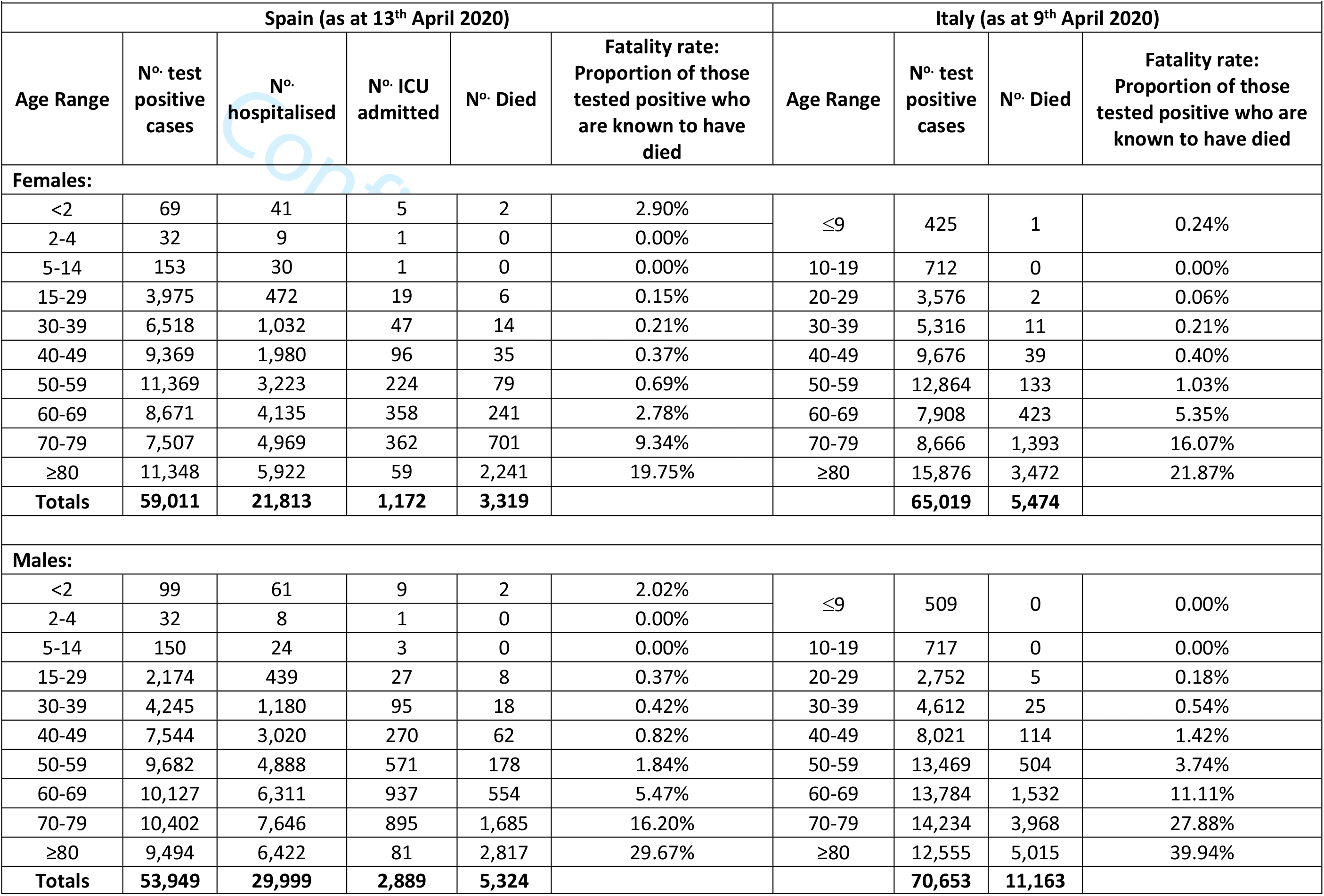
Fatality of health workers from COVID-19 in Spain and Italy as function of sex and age range per positive test rates

|  |  |  |  |  |  |  |  |  |  |
| --- | --- | --- | --- | --- | --- | --- | --- | --- | --- |
| Spain (as at 13<sup>th</sup> April 2020) |  |  |  |  |  | Italy (as at 9<sup>th</sup> April 2020) |  |  |  |
| Age Range | Nº. test positive cases | Nº. hospitalised | Nº. ICU admitted | Nº. Died | Fatality rate: Proportion of those tested positive who are known to have died | Age Range | Nº. test positive cases | Nº. Died | Fatality rate: Proportion of those tested positive who are known to have died |
| Females: |  |  |  |  |  |  |  |  |  |
| <2 | 69 | 41 | 5 | 2 | 2.90% | ≤9 | 425 | 1 | 0.24% |
| 2-4 | 32 | 9 | 1 | 0 | 0.00% |  |  |  |  |
| 5-14 | 153 | 30 | 1 | 0 | 0.00% | 10-19 | 712 | 0 | 0.00% |
| 15-29 | 3,975 | 472 | 19 | 6 | 0.15% | 20-29 | 3,576 | 2 | 0.06% |
| 30-39 | 6,518 | 1,032 | 47 | 14 | 0.21% | 30-39 | 5,316 | 11 | 0.21% |
| 40-49 | 9,369 | 1,980 | 96 | 35 | 0.37% | 40-49 | 9,676 | 39 | 0.40% |
| 50-59 | 11,369 | 3,223 | 224 | 79 | 0.69% | 50-59 | 12,864 | 133 | 1.03% |
| 60-69 | 8,671 | 4,135 | 358 | 241 | 2.78% | 60-69 | 7,908 | 423 | 5.35% |
| 70-79 | 7,507 | 4,969 | 362 | 701 | 9.34% | 70-79 | 8,666 | 1,393 | 16.07% |
| ≥80 | 11,348 | 5,922 | 59 | 2,241 | 19.75% | ≥80 | 15,876 | 3,472 | 21.87% |
| Totals | 59,011 | 21,813 | 1,172 | 3,319 |  |  | 65,019 | 5,474 |  |
| Males: |  |  |  |  |  |  |  |  |  |
| <2 | 99 | 61 | 9 | 2 | 2.02% | ≤9 | 509 | 0 | 0.00% |
| 2-4 | 32 | 8 | 1 | 0 | 0.00% |  |  |  |  |
| 5-14 | 150 | 24 | 3 | 0 | 0.00% | 10-19 | 717 | 0 | 0.00% |
| 15-29 | 2,174 | 439 | 27 | 8 | 0.37% | 20-29 | 2,752 | 5 | 0.18% |
| 30-39 | 4,245 | 1,180 | 95 | 18 | 0.42% | 30-39 | 4,612 | 25 | 0.54% |
| 40-49 | 7,544 | 3,020 | 270 | 62 | 0.82% | 40-49 | 8,021 | 114 | 1.42% |
| 50-59 | 9,682 | 4,888 | 571 | 178 | 1.84% | 50-59 | 13,469 | 504 | 3.74% |
| 60-69 | 10,127 | 6,311 | 937 | 554 | 5.47% | 60-69 | 13,784 | 1,532 | 11.11% |
| 70-79 | 10,402 | 7,646 | 895 | 1,685 | 16.20% | 70-79 | 14,234 | 3,968 | 27.88% |
| ≥80 | 9,494 | 6,422 | 81 | 2,817 | 29.67% | ≥80 | 12,555 | 5,015 | 39.94% |
| Totals | 53,949 | 29,999 | 2,889 | 5,324 |  |  | 70,653 | 11,163 |  |

**Table 2:** Odds Ratio for fatality from data in Table 1

| Table 2: Odds Ratio for fatality from data in Table 1 |  |  |  |  |
| --- | --- | --- | --- | --- |
| Effect |  | Point Estimate | 95% Wald Confidence Limits |  |
| Sex | M vs F | 2.12 | 2.06 | 2.18 |
| Age group 40-49 vs 40-49 |  | 2.15 | 1.65 | 2.815 |
| 50-59 vs 40-49 |  | 5.40 | 4.22 | 6.92 |
| 60-69 vs 40-49 |  | 19.4 | 15.2 | 24.6 |
| 70-79 vs 40-49 |  | 62 | 49 | 79 |
| 80 and above vs 40-49 |  | 115 | 91 | 145 |
| Country | Italy vs Spain | 1.63 | 1.58 | 1.68 |

Information was available for deaths of 92 Italian and xx Spanish doctors. Amongst male doctors, 45 were clinically active aged 65 (62 to 68) (median (25^th^ to 75^th^ centile)), of these 23 were general practitioners, 37 were retired (74 (69 to 80)), 4 between ages of 67 and 81 had returned from retirement to assist in the COVID-19 pandemic. Of 6 women doctors who had died 3 were clinically active (2 GPs and a public health doctor) and aged 57 to 68.

As at 13/04/20 11 male doctors between the ages of 53 and 79 had died (presumed from Covid-19) in the UK, all BAME, median aged 68 (IQR 62 to 76), 16 nurses and midwives had died, 4 men and 12 women, median aged 52 (39 to 58), 43% were from BAME backgrounds. Two hospital porters and a pharmacist had also died.

## Conclusion

The initial crisis management of the Covid-19 pandemic has now moved to a stage of planning in which Human Resources management seeks to be logical and compliant with health and safety legislation. This has included the concept of “Vulnerable Healthcare Workers” who the employer should seek to protect from the excessive risks associated with their predisposition toward mortality or morbidity from exposures in the workplace.

We offer evidence that some staff members are known to be at multiples of risk far higher than those who have been added to the list of “Vulnerable Healthcare Workers”. This evidence will allow employers to add these groups to the list of “Vulnerable Healthcare Workers” who should be protected from direct patient contact with Covid-19.

We suggest that within the working healthcare population, this would apply to males over the age of 50 years and females over the age of 60 years.

The reports of mortality and morbidity available from the health ministries of Spain and Italy are exemplary and the information should be used to optimise the use of staff in the health service and in other industries, in order to preserve life and to reduce the load on hospital beds (especially in ICU) and need for respirators.

### Limitations of this work

The proportion of those who test positive who then die is higher in Italy than in Spain. The protocols for testing were unavailable to us and these may vary from region to region within each of the two countries.

At all ages the proportion of men of those infected who die is twice that of women using our metric. So in each group prioritising women over men when staffing COVID wards should reduce the number falling ill and the need for hospital beds and mortality. The Spanish data includes the number hospitalised and for each age group the proportion of those tested who are hospitalised is lower for women than for men. This may be because of the testing protocols but it would tend to suggest that men get more severe symptoms – maybe more of them have diabetes which is one of the common underlying health conditions in both the Spanish and the Italian reports – in both the proportion of those die who have diabetes is about 30%.

The age gradient is striking – the odds ratio for those 60 to 69 compared to those 30 to 39 is 19. Calls for recently retired doctors and nurses to return to the healthcare systems would need to risk manage this available knowledge.

At the moment when there are still inadequate supplies of Personal Protective Equipment (PPE) managers should try to reduce the risk of losing staff by putting younger doctors and nursing staff onto the COVID-19 wards.

It is not possible to make any statements about ethnicity – although the data from the UK shows that all the doctors and 43% of the nurses who have died had come from a BAME background which would tend to suggest that they are elevated risk of death.

#### “What is already known on this subject?”

COVID-19 is a major pandemic and threat to public health globally. The epidemiology, prevalence, natural history, severity pattern and contact spread and fatality rates are not well known or understood yet these are a major priority from the WHO research priorities for COVID (2). There are anecdotal accounts of the prevalence and impact being different by sex, age and BAME status, however no data are published that add evidence to these accounts

#### “What does this study add?”

This study collates data from public sources for two European countries (Spain and Italy) concerning declared rates of infection and mortality and constructs a preliminary index of fatality for health care workers that shows very large differences in impact between men and women and between young and older people working in healthcare in face to face roles with COVID-19 patients. Men have twice the fatality rates of women and older people (60+) have almost 20 times the fatality rates of young healthworkers (30-39).

**Figure 1:**
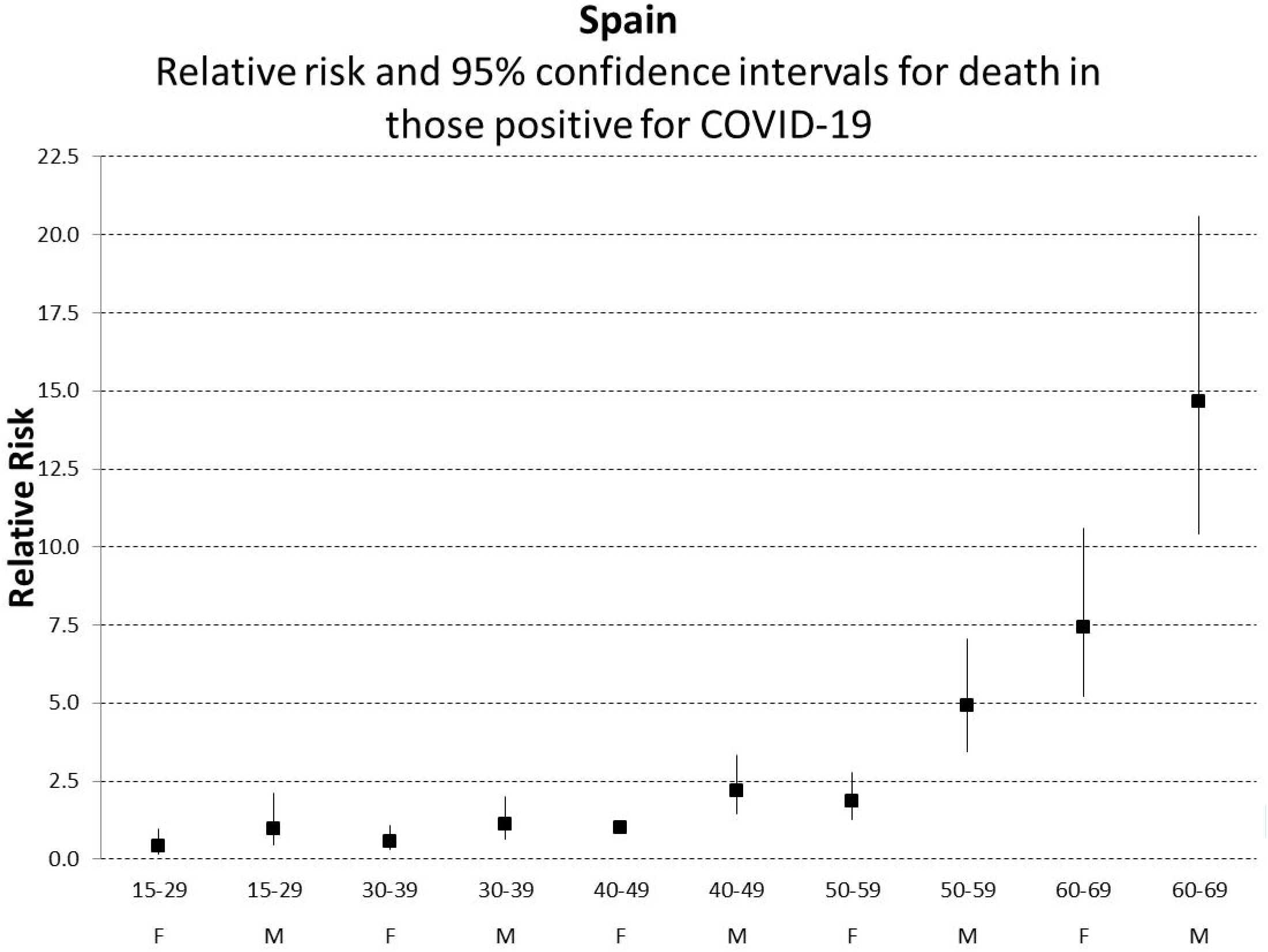
Relative risk and 95% confidence interval for death in Spain, with reference group being women aged 40-49

**Figure 2:**
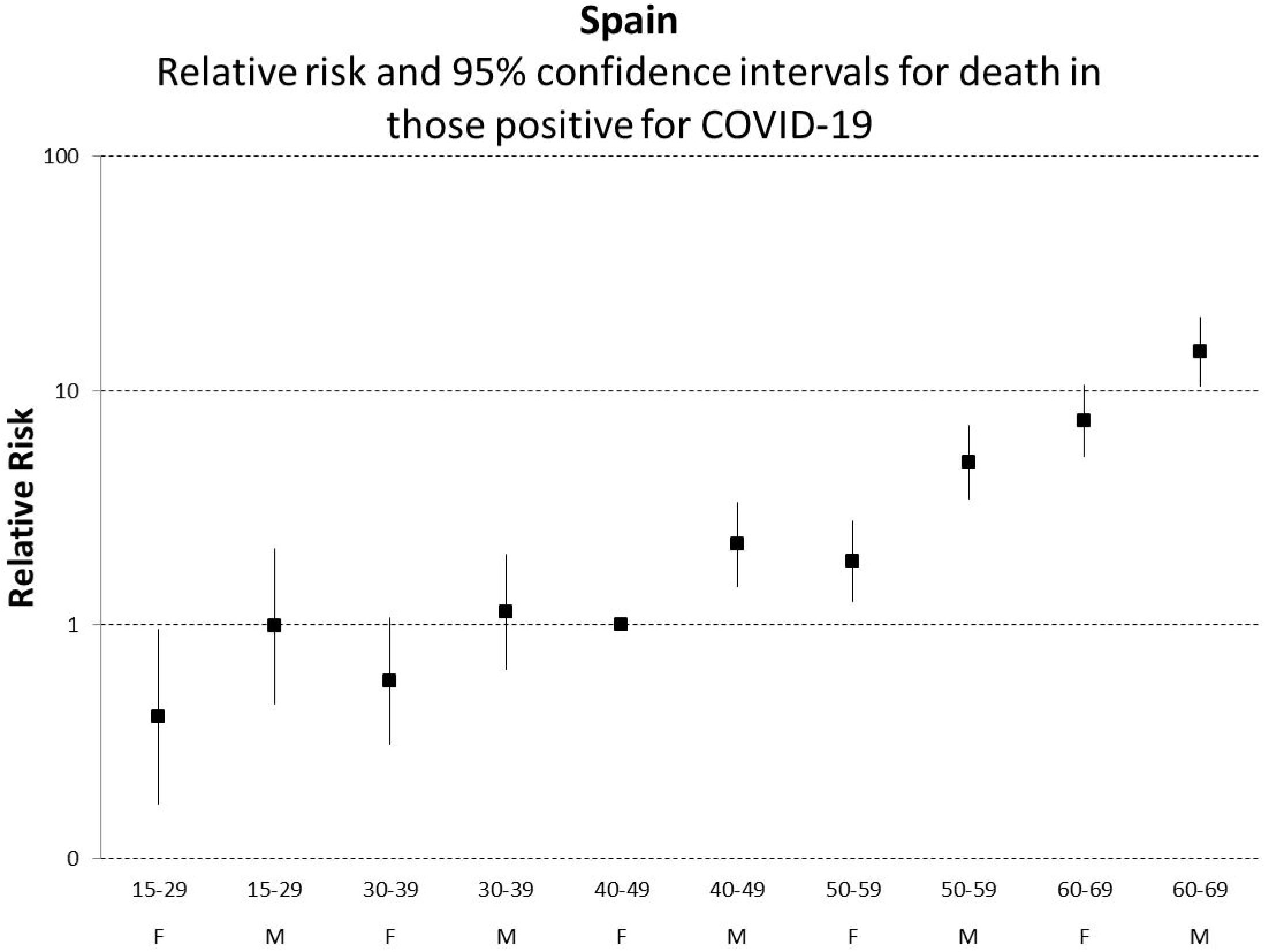
Relative risk and 95% confidence interval for death in Spain, with reference group being women aged 40-49

**Figure 3:**
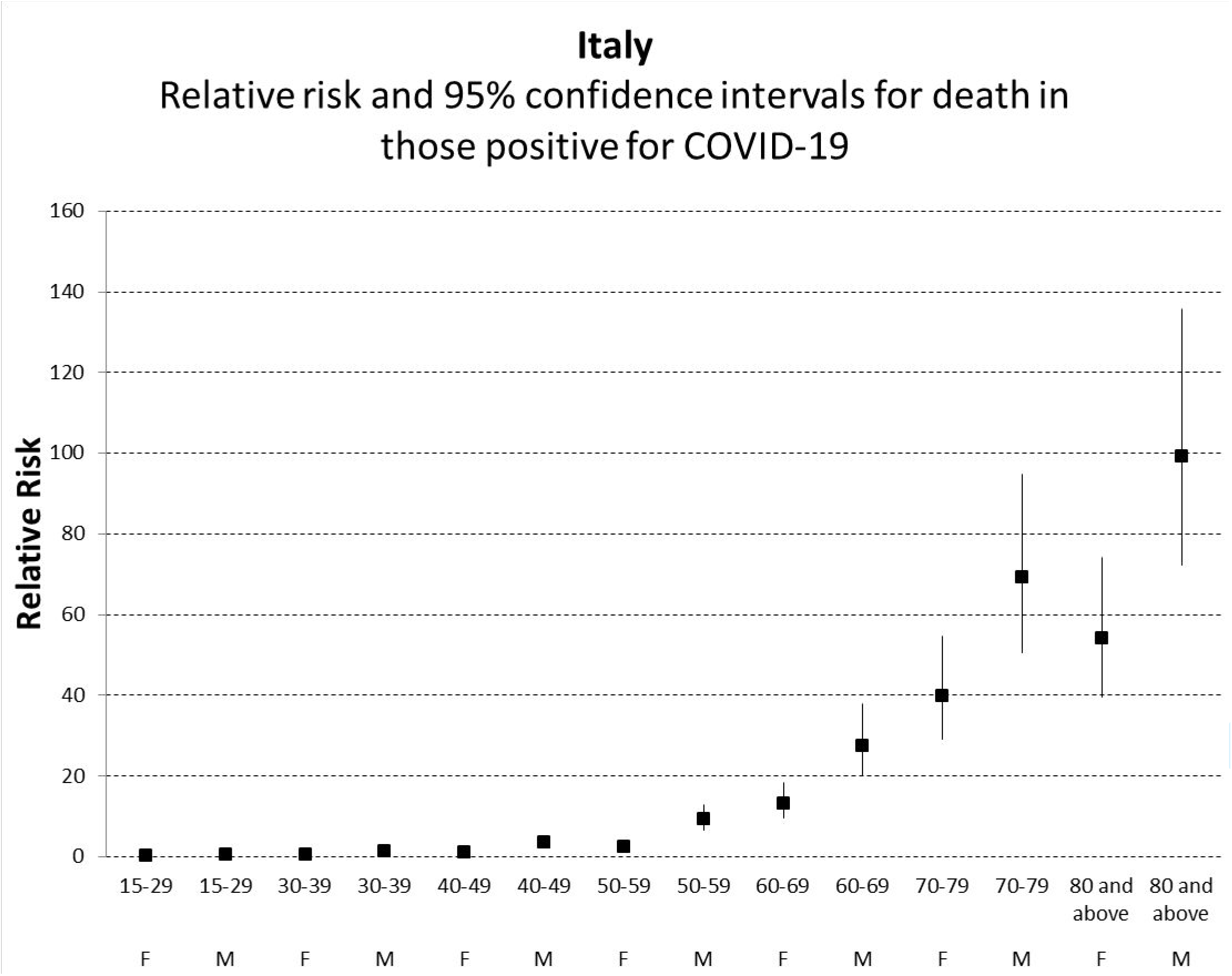
Relative risk and 95% confidence interval for death in Italy, with reference group being women aged 40-49

**Figure 4:**
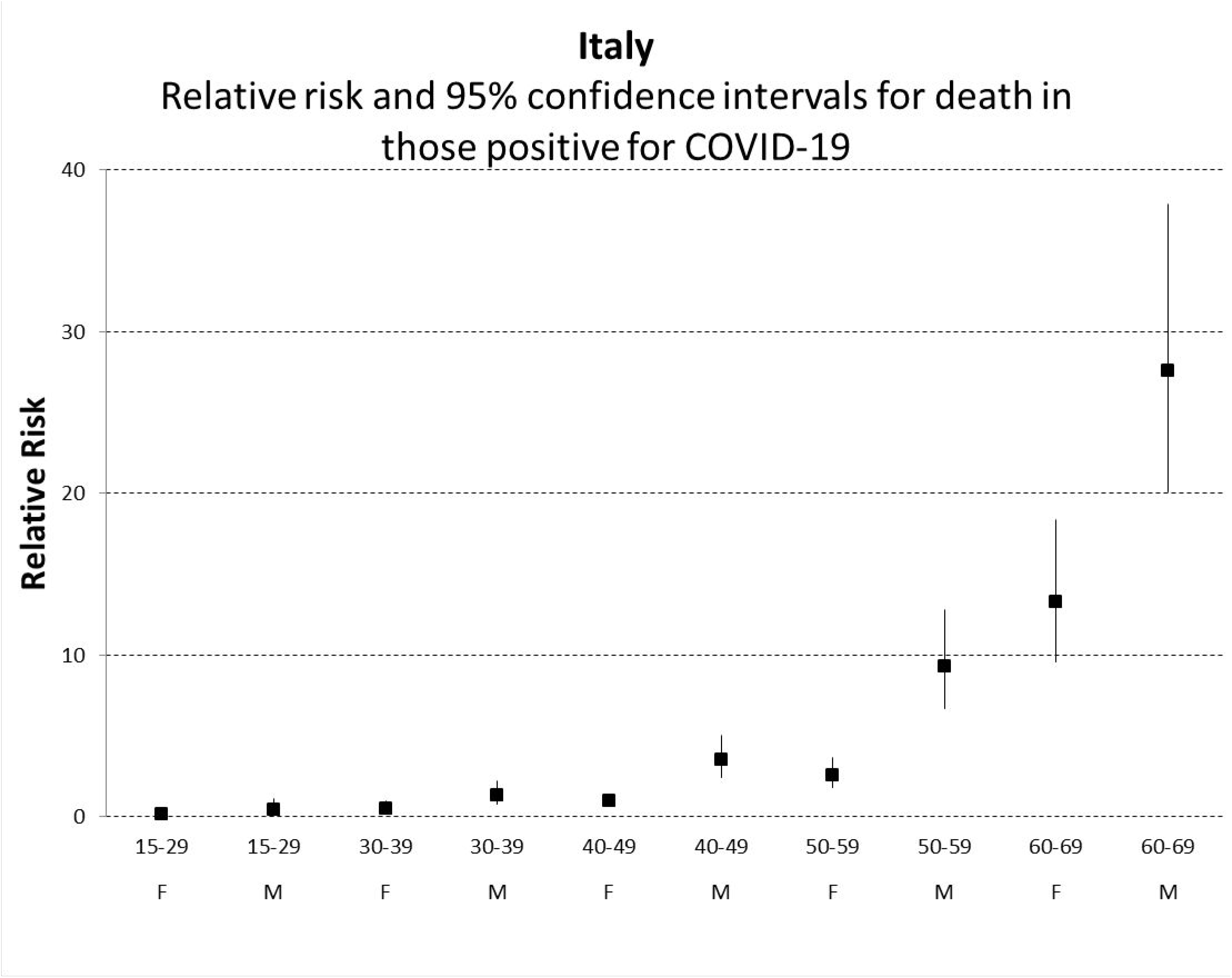
Relative risk and 95% confidence interval for death in Italy, with reference group being women aged 40-49

**Figure 5:**
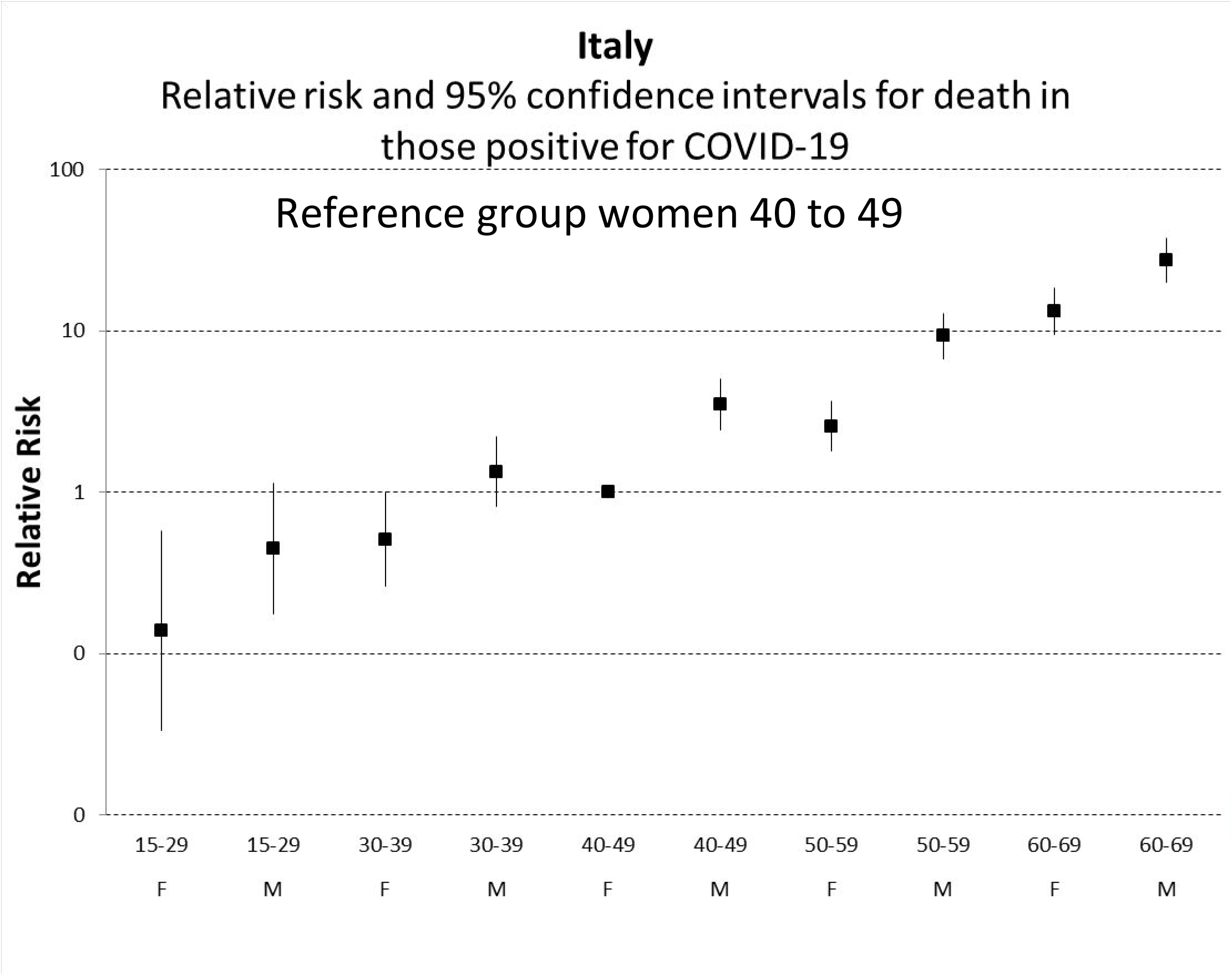
Relative risk and 95% confidence interval for death in Italy, with reference group being women aged 40-49

**Figure 6.**
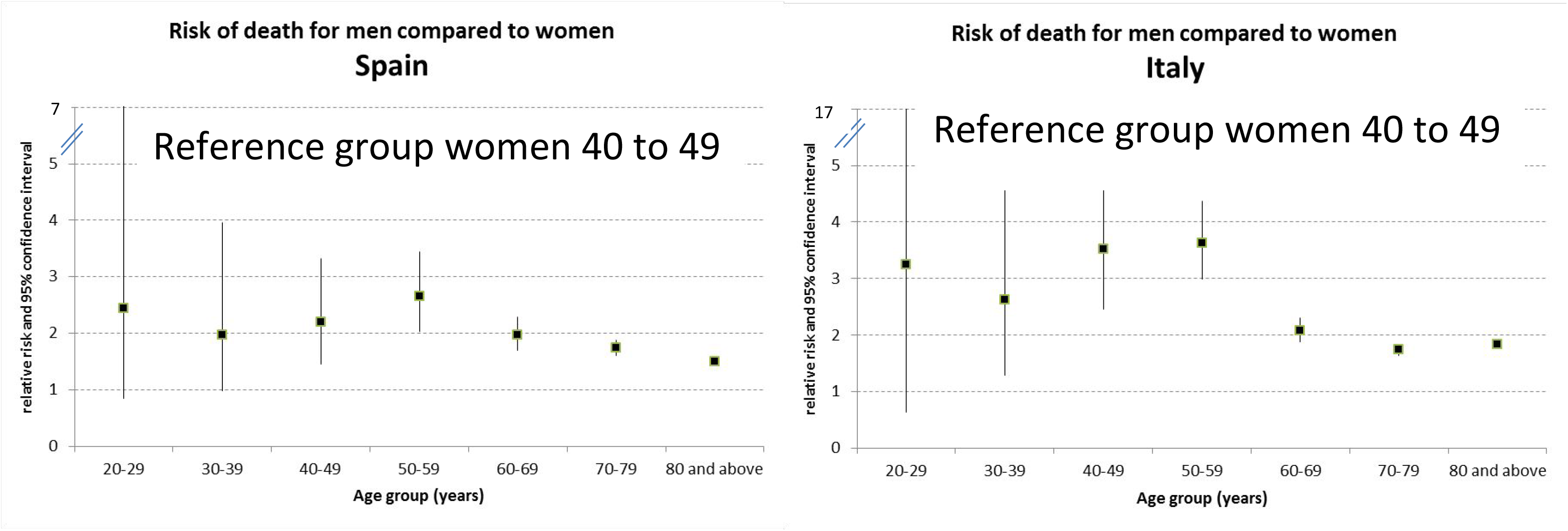
Relative risk and 95% confidence interval for death in men compared to women, taking women aged 40 to 49 as the reference group

## Data Availability

All data are already in the public domain

## References

(1) (https://portale.fnomceo.it/elenco-dei-medici-caduti-nel-corso-dellepidemia-di-covid-19/)

(2) https://www.who.int/who-documents-detail/a-coordinated-global-research-roadmapp15accessed25/4/2018:30

